# Occupational Exposure and Observance of Standard Precautions Among Bucco-dental Health Workers in Referral Hospitals (Yaoundé, Cameroon)

**DOI:** 10.1101/2023.11.24.23298984

**Authors:** Innocent Takougang, Zita Fojuh Mbognou, Fabrice Zobel Lekeumo Cheuyem, Ariane Nouko, Michèle Lowe

## Abstract

**Background:** The risk of infection during dental practices is omnipresent for both patients and healthcare workers. Workers within the bucco-dental health services are among the most affected. The most reported infectious agents transmitted through blood and body fluids are Human Immunodeficiency Virus, viral hepatitis B and C. Compliance with standard precautions prevents exposure to hospital associated infections that are acquired through exposure needle sticks and splashes in healthcare settings. The aim of the present investigation was to assess the level of implementation and constraints to the observance of standard precautions in bucco-dental services.

**Methodology:** A cross-sectional study was conducted in five referral hospitals in Yaoundé, from March to April 2021, involving a purposeful selection of 40 bucco-dental health workers (BDHW). Workers were submitted to a pre-tested self-administered questionnaire covering their knowledge, level of observance of standard precautions and experiences of occupational exposure to blood and other body fluids. The data collected were analyzed using IBM SPSS software version 26.

**Results:** Out of the 53 bucco-dental health workers (BDHW) who were selected for inclusion, 40 provided responses for a participation rate of 75.5%. The mean age of participants was 30.65 years and the M/F sex ratio was 0.54. Half of participants (58.5%) had a good overall level of knowledge of standard precautions. Less than a quarter of participants (12.5%) were compliant with standard precautions. Only 35% of BDHW had received training on hospital infection control, while 60% reported to have experience a needle stick injury in the last three months. Gaps in the observance of standard precautions included the lack of disinfectants (70%), application of hand washing techniques and use of personal protective equipment (PPE). Less than half of participant (47,5%) were fully vaccinated against hepatitis B.

**Conclusion:** Most bucco-dental health workers had insufficient knowledge of standard precautions, most of whom had experienced needlestick injuries and accidental exposure to body fluids, and were at high risk of hospital acquired infections. There is an urgent need to establish and strengthen hospital-based infection control committees to ensure training and implementation of infection prevention measures in local healthcare settings. A framework for the nationwide scale up of such interventions should be explored.

## Introduction

The healthcare sector is the fastest-growing sector of the financial system, employing over 18 million workers worldwide. The hospital environment is one of the most hazardous work settings [1,2]. Healthcare facilities employ some 60 million workers worldwide of whom two-thirds provide health services [3]. Healthcare workers (HCWs) are exposed to biological, chemical, physical and psychosocial hazards, resulting in the loss of some 24 million healthy life years worldwide [4]. HCWs sustain some 3 million percutaneous exposures annually and more than 90% of which occur in resource-limited countries [5,6]. Exposure to healthcare-related hazards is influenced by the occupational category, work experience and environment [7,8]. They cause work-related stress, needlestick injuries, blood and body fluids splashes, and sleep disturbances. Such exposures can lead to hospital acquired infectious agents in body fluids or airborne [9,10]. Potential infectious agents include human acquired immuno-deficiency virus (HIV/AIDS), hepatis viruses A, B and C, airborne tuberculosis and a number of blood-borne pathogens. Some of the acquired infections can be responsible for life-threatening illnesses. Accidental exposure to blood and body fluids (AEBF) result from medical errors, causing more than 20,000 cases of viral hepatitis B, C (HCV) and HIV/AIDS among HCW [11,12].

Accidental exposure to body fluids holds a high risk to the health, safety and quality of care delivered [11]. The working environment, availability of health and safety information, and compliance with standard precautions (SP) are associated with AEBF [12–14]. Key components of SP include hand hygiene, use of personal protective equipment, respiratory protection, development and implementation of related health policies [15]. Adherence to SP aims to minimize occupational exposure to body fluids. Compliance with SP is essential to reduce the spread of infectious agents to patients, HCW and visitors of healthcare settings [16]. Oral health practitioners are disproportionally exposed to healthcare associated infections. The accrued risk is related to the use of rotating instrumentation, aerosols produced during bucco-dental care and the closer contact required with the patient cared for [17]. Dental health practitioners have traditionally been among the HCW reporting the most needle stick injuries [12]. Observance of SP and adherence to vaccination are effective interventions to prevent the transmission of healthcare associated infections [18]. Within the Cameroonian healthcare system, referral hospitals are endowed with providing technical support through continued education including issues related to the promotion of infection control in their satellite establishments [10-13]. The present study aimed to determine the level of compliance with of infection prevention guidelines among dental health workers in referral hospitals.

## Methods

### Study design and period

The present descriptive cross-sectional study was conducted in five purposefully selected referral hospitals, from the March to April 2021. Referral hospitals were the University Teaching Hospital (YUTH), the Central (YCH), Jamot (JHY), Gyneco-Obstetric and Pediatric (YGOPH) and General (YGH). Hospitals. Each of the hospital has a dental unit with cabinets where bucco-dental health workers operate from Monday to Friday. Dental care units typically bear sterilization rooms, prosthesis laboratory, and dental services cabinets with an examination bed, water supply, …

### Study participants

The study participants were dental healthcare workers including doctors, nurses, technicians and assistants, all working in the dental cabinets and accessory services.

### Data collection

Data on infection prevention practices were collected using including a self-administered questionnaire and an observation guide. Observations were made on working days (Monday to Friday) and during daily operating cycle. The questionnaire included topics related to hand hygiene, use of personal protective equipment, respiratory hygiene, injection safety, sharps disposal, antisepsis and sterilization.

### Data processing and analysis

Data collected were entered and analyzed using the SPSS (Statistical Package for the Social Sciences) Version 26. HCWs’ knowledge and practices were assessed with relevance to socio-demographic characteristics. HCW knowledge score were classified as poor (less than 50%), insufficient (50 - 65%), average (65 - 85%) and good (more than 85%). Infection prevention Practices were classified as very bad (less than 50%), bad (50- 65%), inadequate (65-85%), and adequate (>85%).

## Results

Out of 53 bucco-dental healthcare workers (BDHW) contacted, 40 completed the questionnaire, for a response rate of 75.5%.

### Socio-professional characteristics of bucco-dental health workers

Study BDHW were mostly dentists (70%) or dental technicians (22.5%). They were women (65%). Participants aged 20-30 years were the most represented (Table 1).

**Table 1.** Socio-professional Characteristics of Bucco-dental Healthcare Workers in Yaoundé Referral Hospitals, April 2021 (*n=*40)

| Characteristic | Count (n) | Frequency (%) |
| --- | --- | --- |
| **Hospital** |  |  |
| University Teaching | 12 | 30 |
| Central | 14 | 35.0 |
| General | 5 | 12.5 |
| Gyneco-Obstetric and Pediatric | 3 | 7.5 |
| Jamot | 6 | 15 |
| **Sex** |  |  |
| Female | 26 | 65.0 |
| Male | 14 | 35.0 |
| **Age (in Years)** |  |  |
| 20-30 | 30 | 75 |
| 31-40 | 6 | 15 |
| 41 + | 4 | 10 |
| **Professional status** |  |  |
| Dentist | 28 | 70 |
| Dental technician | 9 | 22.5 |
| Dental assistant | 3 | 7.5 |
| **Professional experience (Years of practice)** |  |  |
| < 5 | 26 | 57.5 |
| 5 – 10 | 11 | 27.5 |
| 11 – 20 | 3 | 7.5 |

### Knowledge of Infection prevention and Control

Almost all participants (97.5%) reported being aware of the risk of infection associated with dental care. Half of the participants (50%) had heard of standard precautions. Less than half (35%) had received training in infection prevention in the healthcare setting. The overall level of knowledge of standard precautions as good for more than half (58.5%) (Table 2).

**Table 2.**
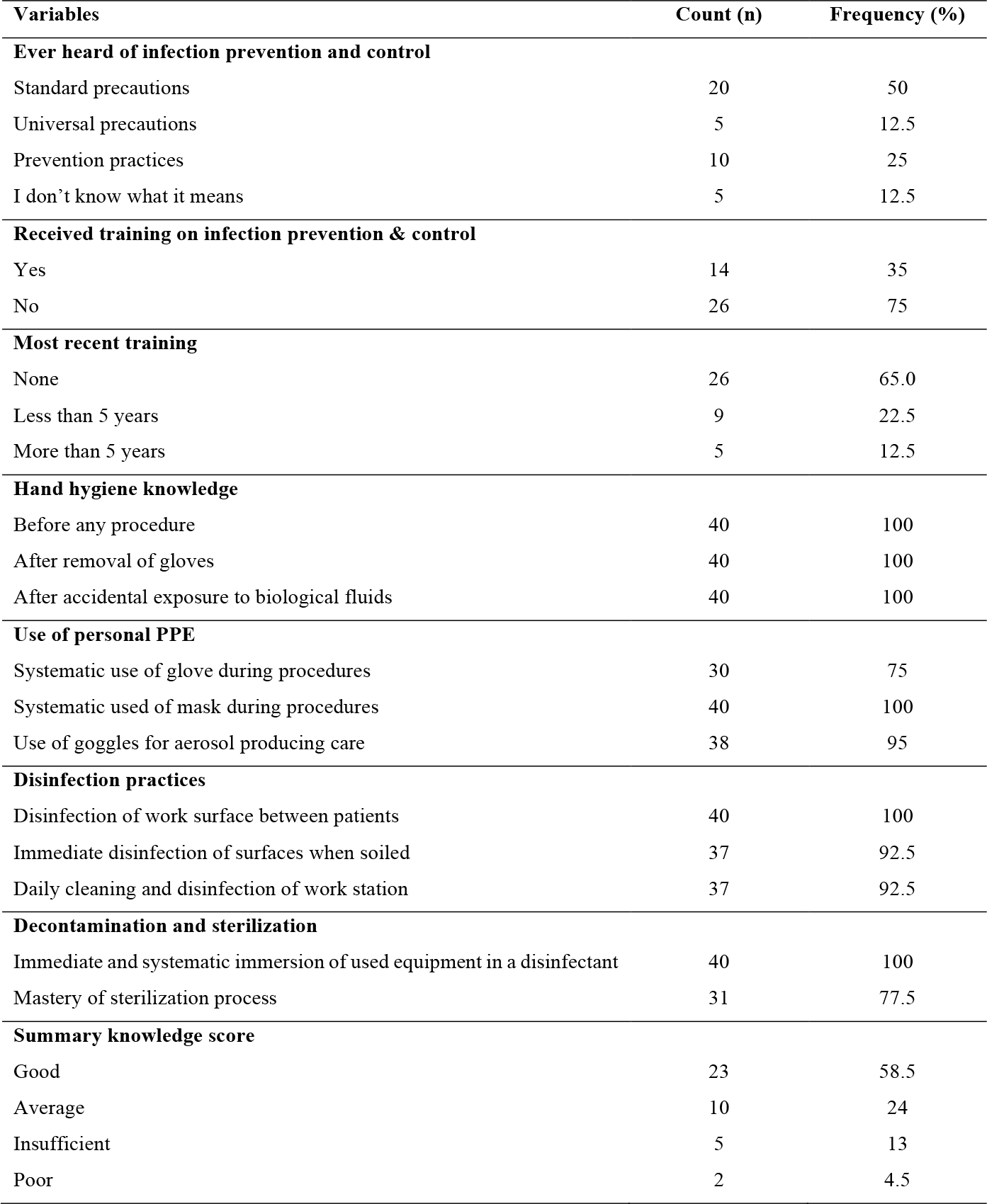
Buco-dental Healthcare Workers’ Knowledge of Standard Precautions in Referral Hospitals in Yaoundé, April 2021 (*n=*40)

| Variables | Count (n) | Frequency (%) |
| --- | --- | --- |
| **Ever heard of infection prevention and control** |  |  |
| Standard precautions | 20 | 50 |
| Universal precautions | 5 | 12.5 |
| Prevention practices | 10 | 25 |
| I don't know what it means | 5 | 12.5 |
| **Received training on infection prevention & control** |  |  |
| Yes | 14 | 35 |
| No | 26 | 75 |
| **Most recent training** |  |  |
| None | 26 | 65.0 |
| Less than 5 years | 9 | 22.5 |
| More than 5 years | 5 | 12.5 |
| **Hand hygiene knowledge** |  |  |
| Before any procedure | 40 | 100 |
| After removal of gloves | 40 | 100 |
| After accidental exposure to biological fluids | 40 | 100 |
| **Use of personal PPE** |  |  |
| Systematic use of glove during procedures | 30 | 75 |
| Systematic used of mask during procedures | 40 | 100 |
| Use of goggles for aerosol producing care | 38 | 95 |
| **Disinfection practices** |  |  |
| Disinfection of work surface between patients | 40 | 100 |
| Immediate disinfection of surfaces when soiled | 37 | 92.5 |
| Daily cleaning and disinfection of work station | 37 | 92.5 |
| **Decontamination and sterilization** |  |  |
| Immediate and systematic immersion of used equipment in a disinfectant | 40 | 100 |
| Mastery of sterilization process | 31 | 77.5 |
| **Summary knowledge score** |  |  |
| Good | 23 | 58.5 |
| Average | 10 | 24 |
| Insufficient | 5 | 13 |
| Poor | 2 | 4.5 |

### Observance of Standard Precautions

Half of the participants reported washing their hands before procedures (50%). Most dental staff reported systematic use of PPE. However, more than half were confronted with PPE shortcut during dental treatment (55%). All participants in our survey worn facial mask including surgical masks (92.5%) and FFP2 masks (7.5%). More than two-thirds of respondents reported disposing single-uses devices correctly (70%). Almost two-thirds of HCW had an inadequate (60%) and detrimental for a quarter of respondents (15%) (Table 3).

**Table 3.** Compliance with Standard Precautions among Dental Care staff in Yaoundé Referral Hospitals, April 2021(*n=*40)

| Standard precaution | Count (n) | Frequency (%) |
| --- | --- | --- |
| **Use of disinfection solution** |  |  |
| Yes | 32 | 80 |
| No | 8 | 20 |
| **Hand hygiene practices** |  |  |
| Short nails and or no nail varnish | 28 | 70 |
| Use of disposable hand towels after hand washing | 11 | 27.5 |
| **Time of hand wash** |  |  |
| Before any procedure | 20 | 50 |
| After any procedure | 40 | 100 |
| Before and after any procedure | 33 | 82.5 |
| After removal of gloves | 30 | 75 |
| **Personal protective equipment** |  |  |
| Use of PPE (gown, cap, clog, over gown, on shoe) | 32 | 80 |
| PPE shortcut (gown, cap, mask, clog, over gown, overshoe) | 22 | 55 |
| Systematic use of goggles/visor | 10 | 25 |
| Systematic use of gloves | 33 | 82.5 |
| Systematic use of facial mask | 40 | 100 |
| Change of gloves between patients | 40 | 100 |
| Change of mask between patients | 8 | 20 |
| Disposal of single-use PPE after use | 12 | 30 |
| **Respiratory hygiene** |  |  |
| Mandatory wearing of masks on entering the dental service | 37 | 92.5 |
| social distancing required between patients | 28 | 70 |
| Isolation of patients with flu-like symptoms | 21 | 52.5 |
| **Waste management and instruments sterilization** |  |  |
| Correct disposal of single-use devices | 28 | 70 |
| Immersion of used equipment in a disinfectant bath | 33 | 82.5 |
| Sterilization in the autoclave at 134°C for 18 minutes | 6 | 15 |
| **Rotating instruments** |  |  |
| Disinfection of rotating instruments | 37 | 92.5 |
| Sterilization of rotating instruments | 5 | 12.5 |
| **Practice score** |  |  |
| Adequate | 5 | 12.5 |
| Inadequate | 24 | 60 |
| Very poor | 5 | 12.5 |
| Detrimental | 6 | 15 |

### Accidental Exposure to Body Fluids

More than half of the respondents (60%) had experience occupational exposure in the last three months. The accidental exposures were related to needlestick injuries. Moreover, less than half of the BDHCW (47,5%) were vaccinated against Hepatitis B. Only one-third of respondents (30%) were knowledgeable about the immediate action recommended following a percutaneous exposure (Table 4).

**Table 4.**
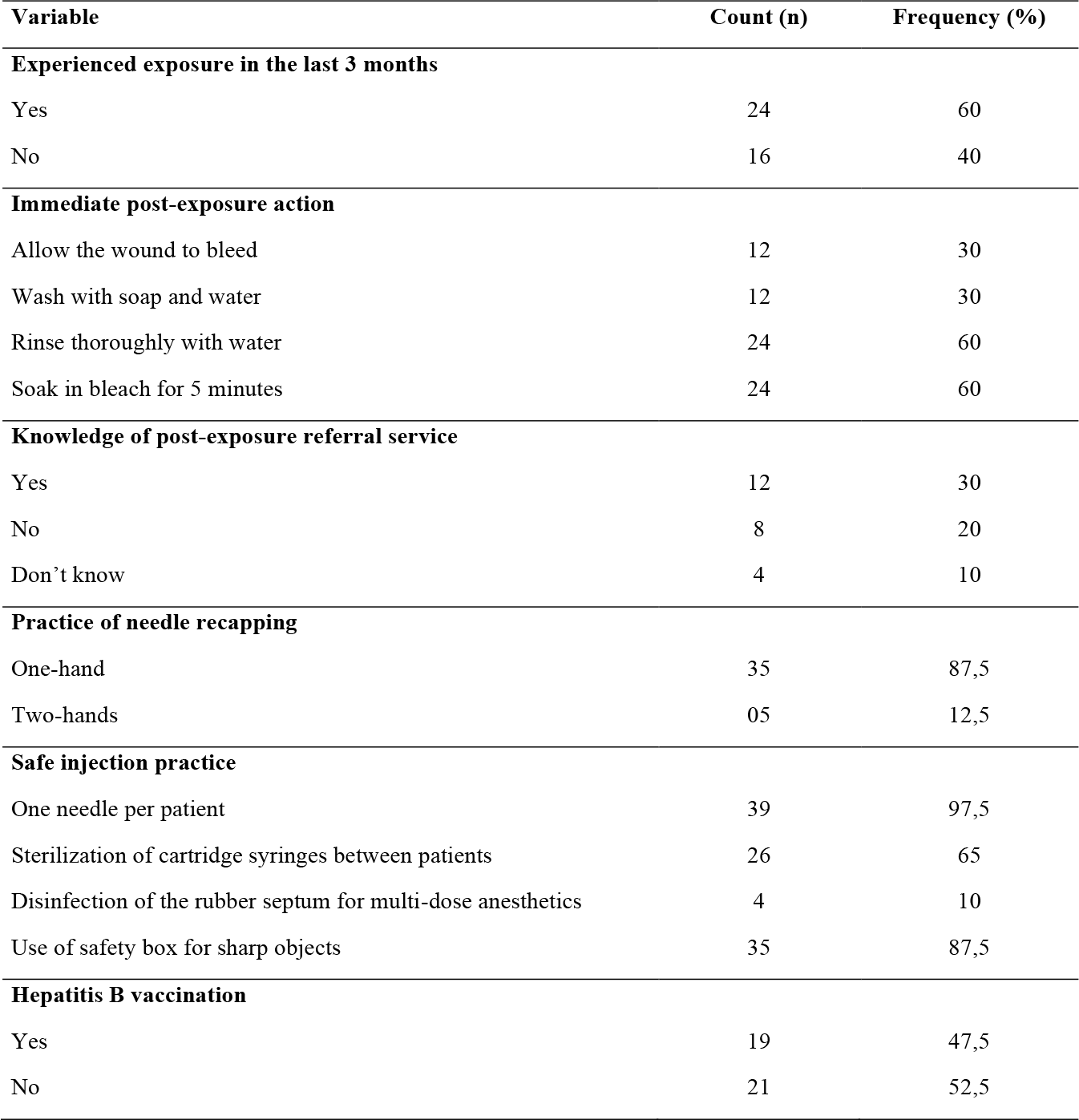
Practices related to Occupational Exposure among Dental Care Staff in Yaoundé Referral Hospitals, April 2021 (*n=*40)

| Variable | Count (n) | Frequency (%) |
| --- | --- | --- |
| **Experienced exposure in the last 3 months** |  |  |
| Yes | 24 | 60 |
| No | 16 | 40 |
| **Immediate post-exposure action** |  |  |
| Allow the wound to bleed | 12 | 30 |
| Wash with soap and water | 12 | 30 |
| Rinse thoroughly with water | 24 | 60 |
| Soak in bleach for 5 minutes | 24 | 60 |
| **Knowledge of post-exposure referral service** |  |  |
| Yes | 12 | 30 |
| No | 8 | 20 |
| Don't know | 4 | 10 |
| **Practice of needle recapping** |  |  |
| One-hand | 35 | 87,5 |
| Two-hands | 05 | 12,5 |
| **Safe injection practice** |  |  |
| One needle per patient | 39 | 97,5 |
| Sterilization of cartridge syringes between patients | 26 | 65 |
| Disinfection of the rubber septum for multi-dose anesthetics | 4 | 10 |
| Use of safety box for sharp objects | 35 | 87,5 |
| **Hepatitis B vaccination** |  |  |
| Yes | 19 | 47,5 |
| No | 21 | 52,5 |

### Constraints to the observance of Standard Precautions

Participants reported the inadequate (40%) or lack of medical supplies, including PPE (90%), as some reasons justifying the low observance of standard precautions. The lack of training sessions on infection prevention in dental care settings was cited as a contributing factor. Some participants (33%) reported the workload and lack of time as justifiers of non-observance of preventive guidelines, while others reported discomfort as a reason for not using the FFP2 masks, face shield and goggles (Figure 1)

**Figure 1.**
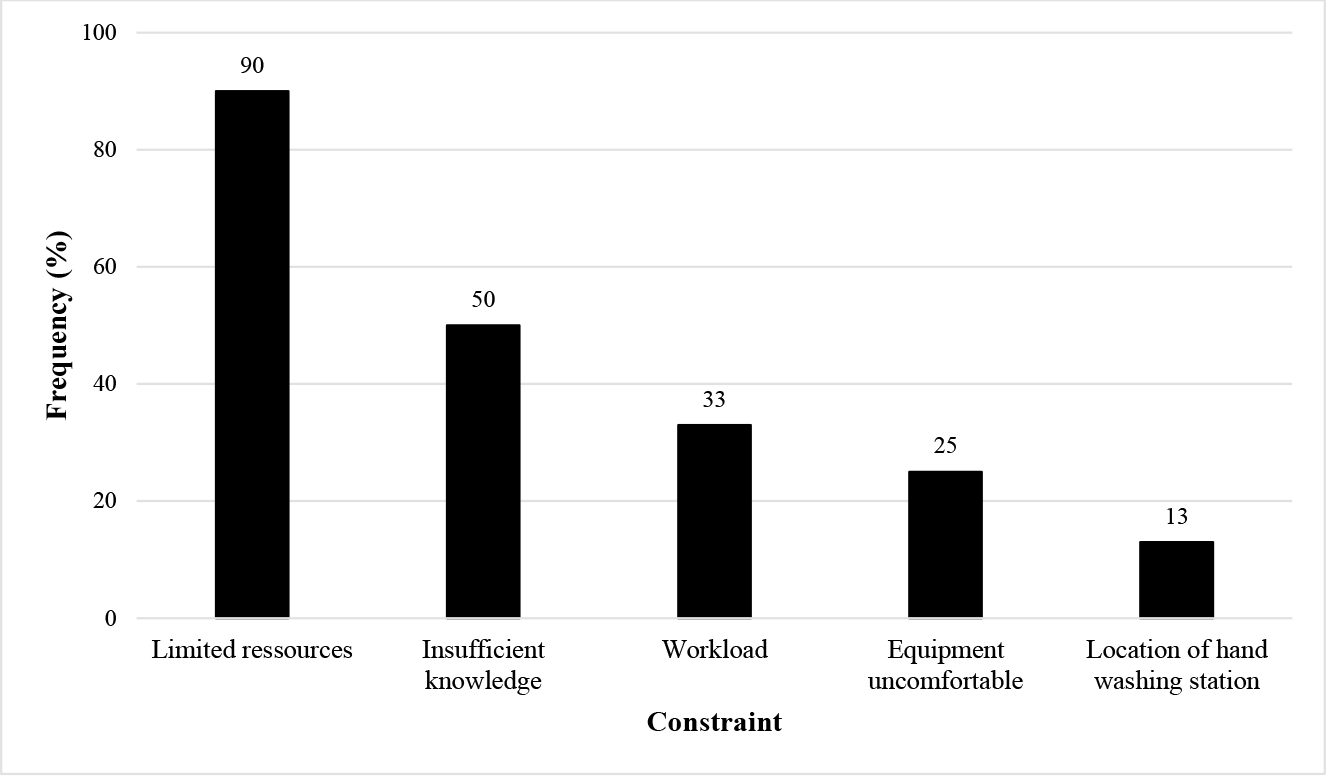
Reported Barriers to the Observance of Standard Precautions among Bucco-dental Health Workers in Yaoundé Referral Hospitals (*n=*40)

## Discussion

The low level of knowledge of standard precautions among BDHW may be related to the inadequacy of the training on the prevention of healthcare-associated infection in the initial curricula and as part of continued education activities. The practice of bucco-dental care requires a closer contact with patients than standard clinical work, exposing to a higher risk of transmission of pathogens. Dental therapeutic procedures are often invasive and produce aerosols that contain biological fluids. All caregivers, patients and the working environment may be subject to soiling. Knowledge and adherence to standard precautions by BDHW is essential to manage their specific health risk associated to exposure to care associated infections.

The research team did not receive confirmation on the existence of infection control committees in the study settings. There is a need to include capacity building of health facilities and health workers at the strategic level, such that both raining/recycling, equipment and supplies to sustain infection prevention activities could be nurtured and prioritized in health policies. Despite the low proportion of trained staff, participants had a good understanding of the practice of hand washing and the systematic use of gloves during bucco-dental care. The low proportion of participants having received SP training in the last five years corroborates observations made elsewhere [21]. There is a need to sustain adherence to good practice through continuing education activities, a compliance booster as BDHW face evolving health technologies and emerging health challenges.

Deficiencies in the supply of PPE, the universal availability of disinfectants, the location of hand washing points were reported as barriers explaining poor compliance with SP among bucco-dental healthcare workers [10].

Though outside the IPC network of activities, heavy workload was reported as a barrier to compliance with standard precautions [22]. Working greater than eight hours per day, working in the night shifts are factors associated with exposure to occupational hazards and medical errors [23].

Almost all participants reported good knowledge and adequate practices related to hand washing. Lower adherence rates (19.4%) were reported in dental care settings in Tunisia [24,25] and Saudi Arabia [26]. The higher adherence reported in the present investigation could be explained by the interest aroused by the COVID-19 pandemic, as there was a widespread availability of disinfectants in healthcare settings. Indeed, the present study took place during the third phase of the COVID-19 pandemic. One of the global measures taken to combat its spread was the promotion and distribution of alcohol-based hand sanitizers in health facilities and public places.

Most participants reported using personal protective equipment, including masks, gloves and gowns when on duty in accordance with the usual instructions [27]. While gloves were routinely worn for all oral and dental care, they were not changed when care was interrupted, a practice similar to those reported in other dental care settings [25–28].

The low rate of use of FFP2 mask was explained by its unavailability in different departments, and its high cost when acquired at the user’s own expense, similar to reports among HCW in Pakistan [27] and Morocco [21].

Less than half of the participants were fully vaccinated against viral hepatitis B. The financial accessibility of the vaccine and the perception of risks could account for the insufficient coverage. Establishing mandatory pre-engagement vaccination against hepatitis B for health workers could be an option [29,30].

The high prevalence of occupational exposure reported in this study is worrying, although it is similar to that reported in Morocco (58.9%) and Côte d’Ivoire (60%) [21,31]. A comprehensive approach including training, awareness raising, implementation of surveillance activities and ensuring adequate supply of necessary equipment and facilities [29,32].

## Conclusion

Half of the bucco-dental healthcare workers had good knowledge of standard precautions. The prevalence of accidental exposure was high and less than half of the BDHW were immunized against hepatitis B. Limited resources, insufficient knowledge and workload were the main reported barriers to the compliance with standard precautions. There is an urgent need to set up infection control committees in the study settings with to ensure occupational safety for HCW optimizing the supply chain of PPE, conducting training, internal monitoring and promoting vaccination.

## Data Availability

All data produced in the present work are contained in the manuscript

## Declaration

### Author’s Contribution

Drafting of the protocol, data collection, analysis and interpretation: ZFM; Drafting of original manuscript: ZFM and AN; Critical revision of the manuscript: FZLC and ML; Conception, design, supervision of implementation, editing and final validation of the manuscript: IT.

### Funding Source

This study received no funding from any agency or organization.

### Ethical Approval Statement

The protocol was approved by Institutional Review Board (IRB) of the Faculty of Medicine and Biomedical Sciences of Yaoundé and the ethical clearance: N°188/UY1/FMS/VDRC/DAASR/CSD issued. Informed consent was obtained from participants prior to inclusion in the study.

### Consent for Publication

Not applicable.

### Declaration of Interests

All authors declare no conflict of interest and approve the final article.

## Acknowledgements

We thank all the health personnel who participated in this study as well as well as the management of the hospitals who gave their authorization for this study to be carried out in their hospitals.

## Notes

### Competing Interest Statement

The authors have declared no competing interest.

### Funding Statement

This study did not receive any funding

### Author Declarations

IRB of the Faculty of Medicine and Biomedical Sciences of Yaounde gave ethical approval for this work

